# Efficacy of PD-(L)1 blockade monotherapy compared to PD-(L)1 blockade plus chemotherapy in first-line PD-L1-positive advanced lung adenocarcinomas: A cohort study

**DOI:** 10.1101/2022.12.05.22283131

**Authors:** Arielle Elkrief, Joao M. Victor Alessi, Biagio Ricciuti, Samantha Brown, Hira Rizvi, Isabel R. Preeshagul, Xinan Wang, Federica Pecci, Alessandro Di Federico, Giuseppe Lamberti, Jacklynn V. Egger, Jamie E. Chaft, Charles M. Rudin, Gregory J. Riely, Mark G. Kris, Marc Ladanyi, Yuan Chen, Matthew D. Hellmann, Ronglai Shen, Mark M. Awad, Adam J. Schoenfeld

## Abstract

**Background:** Single-agent PD-(L)1 blockade (IO) alone or in combination with chemotherapy (Chemotherapy-IO) are approved first-line therapies in patients with advanced lung adenocarcinomas (LUADs) with PD-L1 expression ≥1%. These regimens have not been compared prospectively. The primary objective was to compare first-line efficacies of single-agent IO to Chemotherapy-IO in patients with advanced LUADs. Secondary objectives were to explore if clinical, pathologic, and genomic features were associated with differential response to Chemotherapy-IO vs IO.

**Methods:** This was a multicenter retrospective cohort study. Inclusion criteria were patients with advanced LUADs with tumor PD-L1 ≥1% treated with first-line Chemotherapy-IO or IO. To compare the first-line efficacies of single-agent IO to Chemotherapy-IO, we conducted inverse probability weighted (IPW) Cox proportional hazards models using estimated propensity scores.

**Results:** The cohort analyzed included 874 patients. Relative to IO, Chemotherapy-IO was associated with improved ORR (44% vs 35%, p=0.005) and PFS in patients with tumor PD-L1≥1% (HR 0.75, 95% CI 0.0.61-0.92, p=0.005) or PD-L1≥50% (ORR 55% vs38%, p<0.001; PFS HR 0.74 95%CI 0.56-0.97, p=0.032). Using propensity-adjusted analyses, only never smokers in the PD-L1 ≥50% subgroup derived a differential survival benefit from Chemotherapy-IO vs IO (p=0.03). Among patients with very high tumor PD-L1 expression (≥90%) there were no differences in outcome between treatment groups. No genomic factors conferred differential survival benefit to Chemotherapy-IO vs IO.

**Conclusions:** While the addition of chemotherapy to PD-(L)1 blockade increases the probability of initial response, never-smokers with tumor PD-L1 ≥50% comprise the only population identified that derived an apparent survival benefit with treatment intensification.

## Introduction

Drugs that inhibit PD-1 or PD-L1 (PD-(L)1 blockade) have transformed the treatment landscape for patients with advanced non-small cell lung cancer (NSCLC) without targetable driver mutations. In 2022, there are several US Food and Drug Administration-approved first-line PD-(L)-1 blockade regimens alone (IO) or in combination with platinum-doublet chemotherapy (Chemotherapy-IO), all of which demonstrated clinical benefit compared to chemotherapy^1-7^. The increased breadth of treatment options in newly diagnosed advanced lung cancer and lack of published head-to-head trials makes regimen selection challenging. It is unclear if certain populations might derive similar or greater clinical benefit from IO monotherapy than Chemotherapy-IO, and might thereby avoid the potential toxicities of chemotherapy.

Prior studies have identified clinical and genomic factors associated with efficacy of PD-(L)1 blockade. Eastern Cooperative Oncology Group (ECOG) performance status, smoking history, PD-L1 expression, and tumor mutational burden (TMB) influence clinical activity and outcomes^8-10^. NSCLCs harboring mutations in *STK11* and/or *KEAP1* have worse outcomes to PD-(L)1 blockade, particularly among *KRAS*-mutant NSCLCs^11^. In these cases, addition of chemotherapy might overcome IO resistance^11^. Conversely, other factors such as poor performance status may limit tolerability of chemotherapy and negate potential additive anti-cancer activity of combination therapy^12^.

We posited that clinical, pathologic, and molecular analyses of a real-world patient population could identify patient subpopulations that might differentially benefit from IO vs Chemotherapy-IO in the first-line setting. In order to address biases and differences in baseline characteristics between the IO and Chemotherapy-IO groups, we performed propensity-adjusted analyses in 874 patient cases of lung adenocarcinoma (LUAD) without *EGFR* or *ALK*-sensitizing alterations treated with IO or Chemotherapy-IO at two institutions.

## METHODS

### Patients

After institutional review board approval, patients with advanced LUAD from Memorial Sloan Kettering Cancer Center [MSK] and Dana-Farber Cancer Institute [DFCI] between 2011-2020 were assessed in this retrospective analysis. Only patients with advanced lung adenocarcinoma treated in the first-line with IO or Chemotherapy-IO were eligible for analysis (**Figure S1**).

PD-L1 expression (tumor proportion score) was evaluated in all patients included in the study and reported as the percentage of tumor cells with membranous staining as previously described^13^. For genomic analyses, only patients with genomic sequencing by MSK-IMPACT (MSK) or OncoPanel (DFCI) NGS panels were included^14,15^ (**Supplemental Methods)** TMB was harmonized for comparison as previously published (**Supplemental Methods**)^16^. Patients with tumors with PD-L1 <1% or harboring sensitizing *EGFR* or *ALK* alterations were excluded. Pre-planned analyses in PD-L1 1-49% and ≥50% subgroups were conducted to align with current first-line treatment guidelines^17^. The primary outcomes were objective response rate (ORR), progression-free survival (PFS), and overall survival (OS).

### Statistical Analysis

Descriptive statistics were used to describe the analysis population by treatment group (Chemotherapy-IO vs IO). Differences in baseline characteristics by group were evaluated using the Wilcoxon rank sum test, Fisher’s exact test, or Pearson’s Chi-squared test as appropriate.

Patients who did not experience progression or death by the data lock date (October 21^st^, 2021) were censored at date of last assessment. Investigator-assessed ORR was defined as the rate of partial response + complete response. Real-world investigator-assessed PFS was assessed from the date the patient began therapy to the date of progression as previously described as per RECIST v1.1^18-20^. OS was calculated from treatment start date until date of death or last assessment. Kaplan-Meier curves and log-rank test statistics were computed to compare PFS and OS between groups.

We conducted propensity score modelling for treatment group assignment using a logistic regression model with the following potential prognostic factors as categorical variables: age (≥65 vs <65), Eastern Cooperative Oncology Group performance status (ECOG; scores 2-3 vs scores 0-1), smoking status (current/former smokers vs. never smokers), PD-L1 tumor proportion score percentage (PD-L1 ≥50% vs PD-L1 1-49%), and presence of liver or brain metastases at baseline. To account for the potential difference in treatment assignment, we conducted inverse probability weighted (IPW) Cox proportional hazards models using the estimated propensity scores. Three sets of IPW Cox models were explored. The first model (**main effects model**) contained treatment group along with clinical categorical variables (age, ECOG, smoking status, PD-L1 %, baseline liver metastases and baseline brain metastases) to examine the overall treatment effects on PFS and OS. The second model (**treatment interaction model**) further included interactions between treatment group and clinical variables to examine treatment effect modifications, to identify subgroups for differential benefit of Chemotherapy-IO vs IO. The third model was restricted to patients with genomic data available, building off our second model (**treatment interaction model, genomics analysis cohort**) with the additions of the following categorical variables: harmonized TMB score (see **Supplemental Methods**) and five gene mutations (*KRAS, TP53, KEAP1, STK11*, and *SMARCA4*), as well as their interactions with treatment group. The five gene mutations were selected as they have previously been associated with IO outcomes in lung cancer^21-23^. For each clinical subgroup of interest, we calculated the hazard ratio for treatment group (Chemotherapy-IO vs IO) adjusting for all clinical covariates included in the propensity score IPW Cox proportional hazards model, and visualized these results using forest plots.

Analyses were conducted using R version 4.1.1 with the tidyverse (v1.3.1)^24^, gtsummary (v1.6.0)^25^, survival (v3.3.1) and survminer (v0.4.9) packages^26^.

## Results

### Patient Characteristics

Among all patients with advanced LUAD treated with Chemotherapy-IO or IO without a targetable driver alteration in *EGFR* or *ALK*, 874 patients met criteria for inclusion (**Figure S1**). Median follow-up for the clinical analysis cohort was 23 months (IQR: 12, 38); 397 (45%) patients were treated with Chemotherapy-IO and 477 (55%) were treated with IO (**Table 1**). Relative to the Chemotherapy-IO group, patients in the IO group were slightly older (median age 69 vs 67, p<0.001) with a more frequent and heavier smoking history (median pack years 30 vs. 25 p=0.022) (**Table 1**). PD-L1 high expression (≥50%) was enriched within the IO group relative to the Chemotherapy-IO group (84% vs 29% with PD-L1 high, p<0.001) (**Figure S2A**)^17^. Median harmonized TMB score (0.09 vs -0.05, p=0.023) was also significantly higher in the IO group (**Figure S2B)** Other baseline clinical factors such as ECOG performance status and sex were similarly distributed between the Chemotherapy-IO and IO groups (**Table 1**).

### Clinical Outcomes

Among 874 patients with PD-L1 ≥1% LUAD who were treated with Chemotherapy-IO or IO in the first-line setting, Chemotherapy-IO was associated with improved ORR (44% vs 35%, p=0.005) (**Figure 1A**) and improved PFS (median PFS 6.9 (95% CI: 6.0, 8.5) vs 4.9 months (95% CI: 4.0, 5.8), p=0.015) (**Figure 1B**). There was no difference in OS between the Chemotherapy-IO vs IO groups (median OS 17 months, 95% CI: 15, 22) vs 20 months (95% CI: 17, 24), p=0.50) (**Figure 1C**). Examining subgroups of interest, age <65 (HR 0.59, 95%CI 0.42-0.8), and never smoking status (0.43 95%CI 0.22-0.83) were particularly associated with improved PFS among patients on Chemotherapy-IO (**Figure 1D**). No subgroup conferred differential survival benefit to Chemotherapy-IO vs IO (**Figure 1E**).

**Figure 1.**
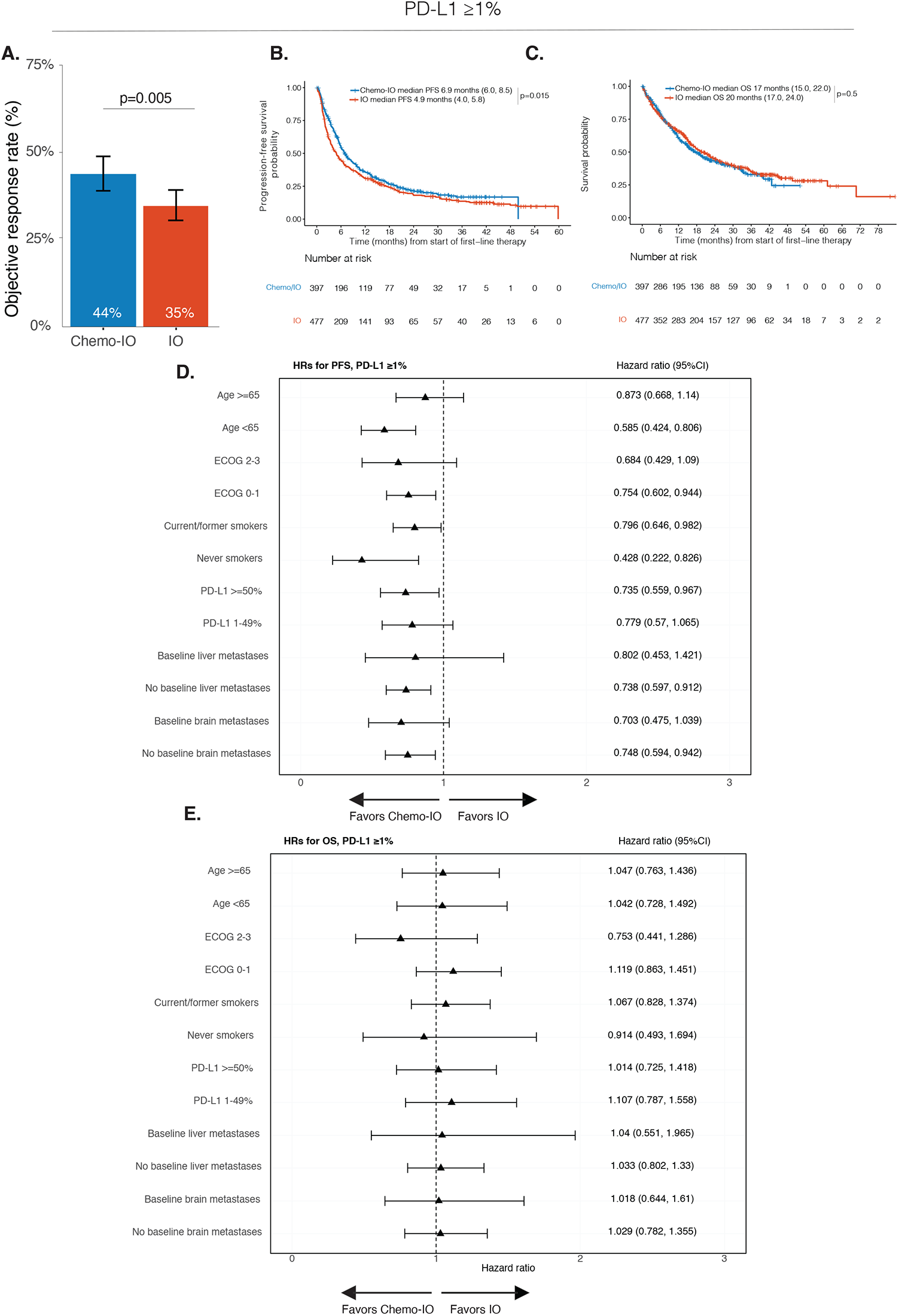
Comparative effectiveness of chemotherapy plus PD-(L)1 blockade (Chemotherapy-IO) vs single-agent PD-(L)1 blockade (IO) in patients with PD-L1≥1%. **A**. Objective response rate, **B**. Progression-free survival (PFS), and **C**. Overall survival (OS) among patients with tumor PD-L1≥1% who received first-line Chemotherapy-IO (N=397) vs IO (N=477). **D**. PFS and **E**. OS analysis for Chemotherapy-IO vs IO adjusting for co-variates of interest among different subgroups. Median survival times presented with confidence intervals in brackets. ECOG; Eastern Cooperative Oncology Group performance status. HR; hazard ratio. 95%CI; 95% confidence intervals.

In the IPW propensity-adjusted analysis, current/former smokers (OR 0.48, 95%CI 0.28-0.79, p=0.004) those with tumor PD-L1 ≥50% (OR 0.07, 95% CI 0.05-0.10, p<0.001) were more likely to receive IO than Chemotherapy-IO (**Table S1**). The results for the main effects model are shown in **Table S2 and Table S3**. In the treatment interaction model, Chemotherapy-IO was associated with superior PFS compared to IO alone (HR 0.38, 95% CI 0.17-0.86, p=0.020) (**Table S4**). No subgroups demonstrated differential PFS benefit for Chemotherapy-IO or IO (**Table S4**).). There was no OS benefit associated with Chemotherapy-IO (HR 0.92, 95% CI 0.42-2.00, p=0.824) (**Table S5**). No subgroups demonstrated differential OS benefit to Chemotherapy-IO or IO (**Table S5**).). In sum, among patients with PD-L1 ≥1%, Chemotherapy-IO demonstrated a PFS benefit in a propensity-matched analysis; however, there was no OS benefit associated with Chemotherapy-IO compared to IO monotherapy.

### Clinical Outcomes by PD-L1 Subgroup

Among 519 patients with tumor PD-L1 ≥50%, Chemotherapy-IO was associated with improved ORR (55% vs 38%, p<0.001) (**Figure 2A**) and PFS (median PFS 10.0 (95% CI: 6.9, 16.0) vs 4.9 months (95% CI: 3.9, 6.2), p=0.002) (**Figure 2B**). There was no significant difference in OS between the Chemotherapy-IO vs IO groups (median OS 32 (95% CI: 20, not reached (NR)) vs 20 months (95% CI: 17, 26), p=0.40) (**Figure 2C**). Examining subgroups of interest, never-smokers (HR 0.29, 95%CI 0.16, 0.55) and younger patients <65 years of age (HR 0.47 95%CI 0.31, 0.71), had improved PFS to Chemotherapy-IO vs IO (**Figure 2D**).

**Figure 2.**
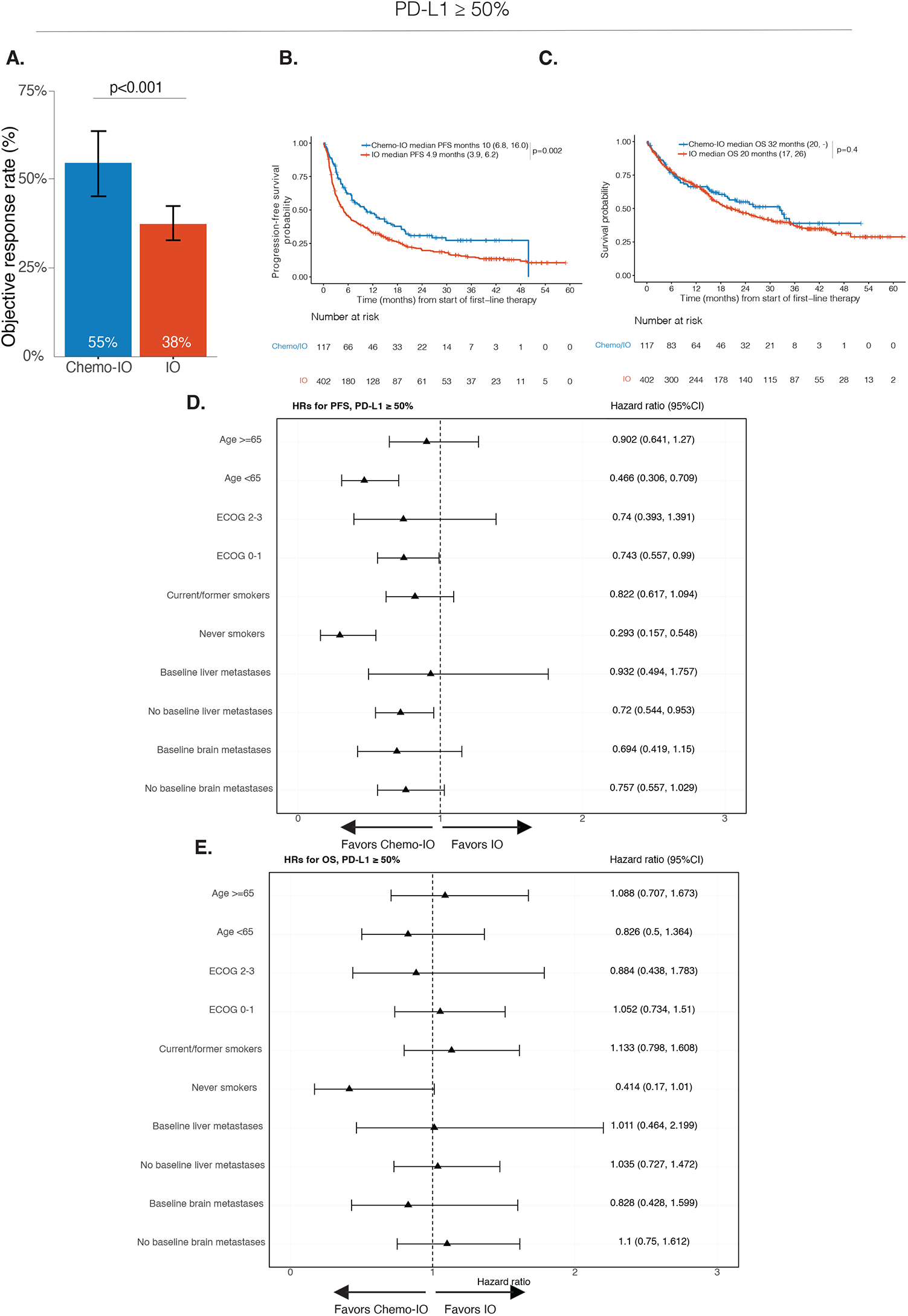
Comparative effectiveness of chemotherapy plus PD-(L)1 blockade (Chemotherapy-IO) vs single-agent PD-(L)1 blockade (IO) in patients with PD-L1≥50%. **A**. Objective response rate, **B**. Progression-free survival and **C**. Overall survival among patients with tumor PD-L1≥50% who received first-line Chemotherapy-IO (N=117) vs IO (N=402). **D**. PFS and **E**. OS analysis for Chemotherapy-IO vs IO adjusting for co-variates of interest among different subgroups. Median survival times presented with confidence intervals in brackets. ECOG; Eastern Cooperative Oncology Group performance status. HR; hazard ratio. 95%CI; 95% confidence intervals.

In the IPW propensity-adjusted analysis, patients who were younger (<65 years of age) (HR 0.58, 95%CI 0.38-0.90, p=0.015) or who were never-smokers (HR 0.52, 95% CI 0.28-1.01, p=0.045) were more likely to receive Chemotherapy-IO (**Table S6**). The results for the main effects model are shown in **Table S7 and Table S8**.

We found significant treatment effect modification factors shown in the treatment interaction model in this PD-L1≥50% subgroup (**Table S9**) Specifically, the Chemotherapy-IO PFS benefit over IO alone was more evident in never-smokers (p=0.0089) (**Table S9**). Age was identified as a significant treatment modifier as well (p=0.0325, **Table S9**). Specifically, the Chemotherapy-IO benefit over IO alone was more evident in the younger age group (<65 years) (**Table S9**). There was no significant difference in OS for Chemotherapy-IO vs IO overall (HR 0.36, 95%CI 0.12-1.10, p=0.0718) (**Table S10**). Never-smokers derived an OS benefit to Chemotherapy-IO vs IO (HR 2.98, 95%CI 1.08-7.9, p=0.034) (**Table S10**). Examining clinical outcomes in these subgroups in greater detail, ORR in never-smokers was higher with Chemotherapy-IO vs IO group (76% vs 20%, p<0.001, respectively) (**Figure 3A**). Median PFS among never-smokers in the Chemotherapy-IO vs IO group was 10.0 months vs 2.2 months, respectively (**Figure 3B**). ORR among patients <65 years of age in the Chemotherapy-IO vs IO groups was higher (63% vs 40%, respectively; p=0.004) (**Figure 3D**). Median PFS among <65 years of age in the Chemotherapy-IO vs IO group was 17.0 months vs 4.6 months, respectively (**Figure 3E**). Taken together, these results suggest that in the PD-L1 ≥50% subgroup, there was a PFS benefit in never-smokers and in younger adults, and OS benefit in never-smokers favoring Chemotherapy-IO compared to IO.

**Figure 3.**
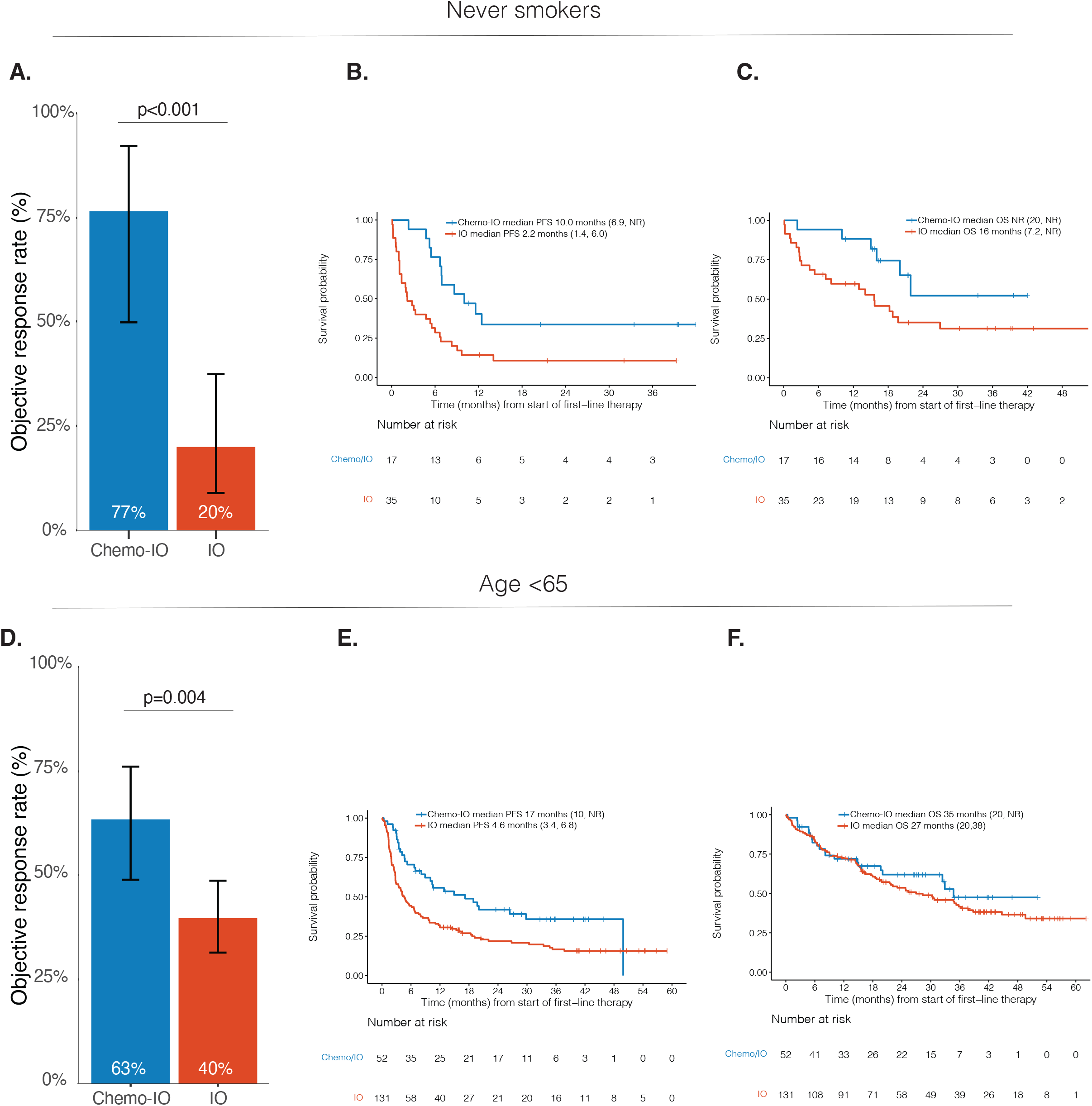
Comparative effectiveness of chemotherapy plus PD-(L)1 blockade (Chemotherapy-IO) vs single-agent PD-(L)1 blockade (IO) in patients with PD-L1≥50% in subgroups of interest. **A**. Objective response rate, **B**. Progression-free survival (PFS), and **C**. Overall survival (OS) between Chemotherapy-IO vs IO groups in never smokers with tumor PD-L1≥50%. **D**. Objective response rate, **E**. PFS, and **F**. OS between Chemotherapy-IO vs IO groups in patients <65 years of age with PD-L1≥50%. Median survival times presented with confidence intervals in brackets.

Next, since very high PD-L1% (≥90%) has previously been associated with improved outcomes to IO^27^, we explored this group in our dataset. Among patients with very high tumor PD-L1 expression (≥90%, N=214) there was no difference in ORR (p=0.2) (**Figure S3A**), PFS (p=0.15) (**Figure S3B**) or OS (p=0.4) (**Figure S3B**) between the Chemotherapy-IO or IO groups.

Lastly, in the PD-L1 1-49% subgroup (N=355 patients), Chemotherapy-IO was associated with improved ORR (39% vs 19%, p<0.001) (**Figure S4A**) and PFS (median PFS 6.2 (95% CI: 5.5, 7.5) vs 4.4 months (95% CI: 3.4, 6.7), p=0.016) (**Figure S4B**) compaed to IO. There was no difference in OS between the Chemotherapy-IO vs IO groups (median OS 15 (95% CI: 13, 19) vs 17 months (95% CI: 10, 24), p=0.7) (**Figure S4C**). IPW propensity-adjusted analysis and treatment interaction models for the PD-L1 subgroup are presented in **Tables S11-S15;** there was no differential benefit to Chemotherapy-IO compared to IO in this subgroup for PFS or OS in any subgroup examined.

### Differential genomic biomarkers of response to Chemotherapy-IO vs IO

There were 579 patients with tumor PD-L1 ≥1% who were treated with Chemotherapy-IO (N=263) or IO (N=316) and underwent genomic sequencing (**Table S1**, genomic analysis cohort). Median follow-up for the genomic analysis cohort was 24 months (IQR: 13, 40). Patient baseline characteristics for the genomic analysis cohort were similar to the clinical analysis cohort (**Table S16**).

There were no differences in ORR between Chemotherapy-IO and IO within different mutation subgroups of interest in *KRAS, TP53, SMARCA4, KEAP1, STK11, or* TMB (**Figure 4A**). In propensity-adjusted analyses, significant treatment effect modifiers were found. Specifically, the Chemotherapy-IO benefit over IO alone was more evident in patients with higher harmonized TMB score (p=0.002) (**Table S17**). Conversely, the Chemotherapy-IO benefit over IO alone was reduced in patients with *SMARCA4* mutations (p=0.011) (**Table S17**). No significant differential OS benefit to Chemotherapy-IO vs IO was found among patients with mutations in *KRAS, KEAP1, STK11*, or *TP53* or among TMB thresholds (**Table S18**). Examining the significant covariates conferring differential initial benefit to Chemotherapy-IO vs IO further, there were no differences in PFS or OS among patients with either high or low median TMB (**Figure 4B-E**). Patients with *SMARCA4* had numerically improved median PFS (**Figure 4F**) and OS (**Figure 4G**) to IO compared to Chemotherapy-IO.

**Figure 4.**
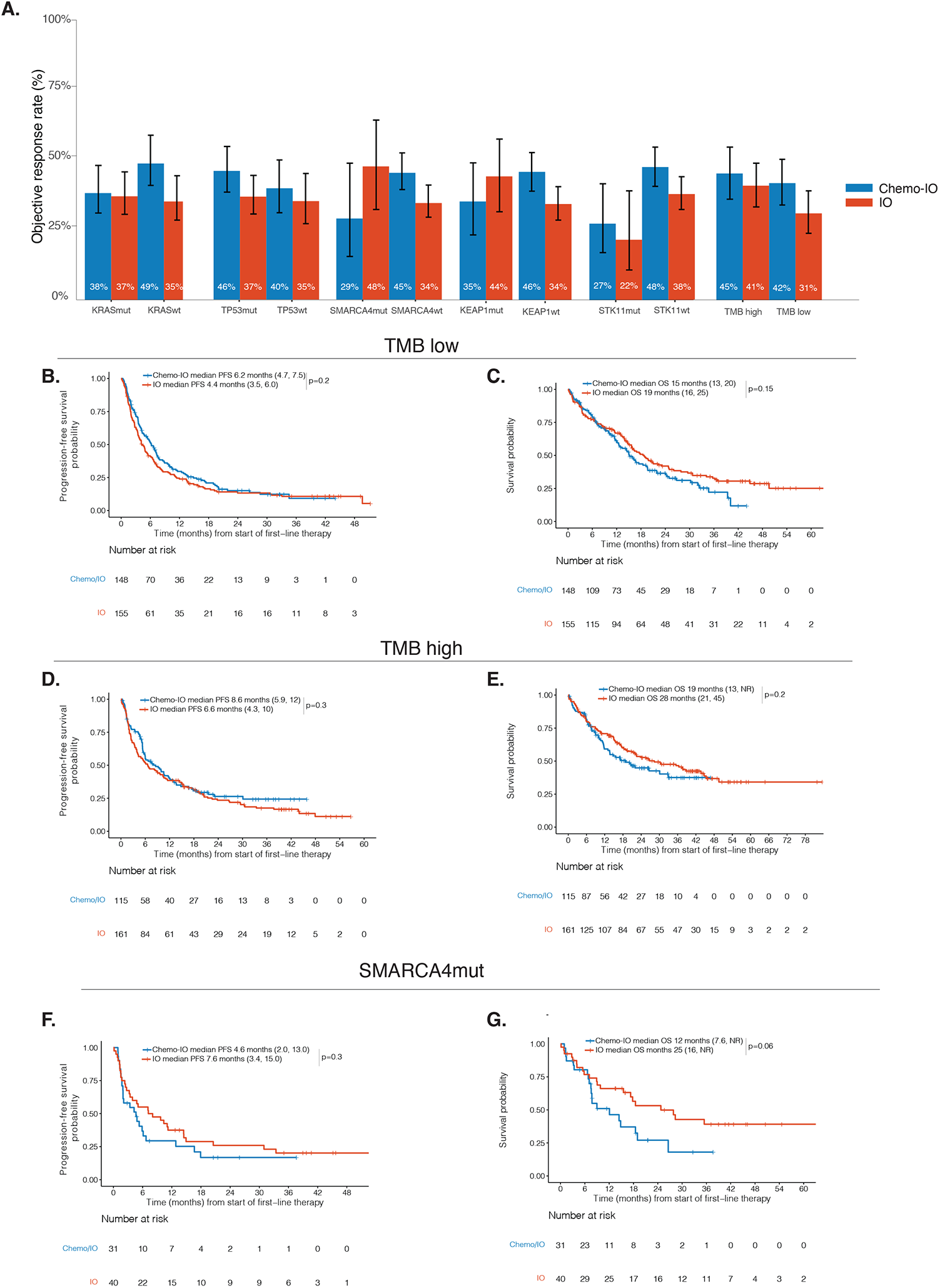
Comparative effectiveness of chemotherapy plus PD-(L)1 blockade (Chemotherapy-IO) vs single-agent PD-(L)1 blockade (IO) in the genomic analysis cohort (PD-L1≥1%). **A**. Objective response rate among genomic subgroups of interest (*KRAS, TP53, SMARCA4, KEAP1, STK11, TMB)*. **B**. Progression-free survival (PFS) and **C**. Overall survival (OS) of Chemotherapy-IO vs IO in TMB low groups. **D**. PFS and **E**. OS for the TMB high groups. **F**. PFS and **G**. OS of Chemotherapy-IO vs IO in patients with *SMARCA4* mutations. Median survival times presented with confidence intervals in brackets. TMB; tumor mutational burden. TMB high; TMB >=50^th^ percentile, TMB low; TMB <50^th^ percentile. Mut; mutated. WT; wild type. Mut; mutated. WT; wild type. Median survival times presented with confidence intervals in brackets.

### Discussion and conclusion

We examined the efficacy of IO compared to treatment intensification with Chemotherapy-IO, in the context of key clinical and molecular features of LUAD. Within the PD-L1 ≥50% subgroup, we found the addition of chemotherapy to PD-(L)1 blockade led to improvements in ORR and PFS, but this only translated into an OS benefit in never-smokers. The only population that did not derive any initial benefit to Chemotherapy-IO over IO was the PD-L1 very high (≥90%) subgroup.

Our analyses inform potential rational strategies to select and sequence available therapies for patients with advanced lung cancers without targetable driver mutations. The lack of superior survival with Chemotherapy-IO compared to IO supports the notion that, biologically, chemotherapy and immunotherapy may be working independently and additively rather than generating synergistic immunologic activity^28^. The addition of chemotherapy increases the probability of initial response in a heterogenous patient population with differential sensitivity to chemotherapy and immunotherapy, but long-term benefit appears largely driven by whether PD-(L)1 blockade generates durable anti-tumor immunity. The lack of OS benefit with Chemotherapy-IO compared to IO in our overall study population is also supported by recent clinical analyses of real-world and randomized-controlled trials in patients with PD-L1 ≥50% non-squamous NSCLC ^18,29^. In these circumstances, deeper investigation of clinical and genomic factors associated with differential benefit with Chemotherapy-IO compared to IO is helpful to optimize regimen selection.

We initially hypothesized that distinct clinical and molecular characteristics previously associated with IO resistance (for example, baseline liver metastases, *STK11*, and/or *KEAP1* mutations) might distinguish patient populations that would benefit from treatment intensification with Chemotherapy-IO ^11^. Contrary to our hypothesis, we found that while factors such as liver metastases and *STK11* mutations were associated with poor response overall, they did not confer superior long-term benefit from Chemotherapy-IO. Thus, treatment escalation with chemotherapy may not be sufficient or indicated solely based upon these factors. Conversely, predictors of durable benefit from IO such as very high PD-L1% (≥90%) may be a useful indicators of when IO monotherapy is appropriate^30^. For this select group of patients, we found no difference in initial response between the Chemotherapy-IO or IO groups. Interestingly, our study also found that patients with *SMARCA4* mutations experienced better response rates and PFS with IO compared to Chemotherapy-IO. The clinical implication of this observiation is supported by prior studies, which have identified *SMARCA4* mutation as a poor prognostic factor, potentially associated with resistance to chemotherapy^31,32^ but improved response to IO [17]. However, only 12% of patients had *SMARCA4* mutations. More extensive molecular and gene expression analyses may be helpful in determining if specific genomic signatures benefit differentially from Chemotherapy-IO vs IO, but these analyses require much larger sample size for adequate power in the face of multiple hypothesis testing.

Never smoking status emerged as the primary factor associated with survival benefit favoring Chemotherapy-IO within the PD-L1 ≥50% subgroup. Prior studies have found a correlation between smoking history and increased TMB and that smoking history could be a potential clinical surrogate for TMB^33,34^. However, here we found that even when accounting for TMB in a propensity adjusted model, smoking status remained a distinct predictive factor. Smoking history is a readily available biomarker that could be of value in determining regimen selection. Our analysis suggests that Chemotherapy-IO should be strongly considered for never-smokers even in the presence of high TMB and/or PD-L1 expression. Further work is needed to identify the underlying biologic basis for this pattern.

The increased response rate and PFS benefit generally observed with Chemotherapy-IO suggests that therapeutic escalation could have an important role in aggressive or extensive disease. Here, the addition of chemotherapy could help provide more immediate symptom relief. Further, these patients may not otherwise have the opportunity to receive salvage chemotherapy after progression on immunotherapy. In this scenario, initial disease control with chemotherapy could create a window of disease control to enable the more gradual effects of IO to become operant. Notably, however, in the setting of brain or liver metastases, we did not find any clinical benefit from the addition of chemotherapy to IO, consistent with prior analysis^35^. In this setting, it is unclear if the added toxicity that comes with chemotherapy may abrogate the potential clinical benefits. Further investigation is also needed to understand the precise impact of volume of disease on patient outcomes.

Our study is a retrospective analysis with the associated limitations, but we sought to minimize inherent bias by including two large institutions and by applying robust propensity-matching methodology. The modest sample size of the PD-L1 1-49% subgroup, especially within the patients who received single-agent IO limited the power of the analysis for this sub-population. Despite these limitations, our study is the first to comprehensively study the comparative effectiveness of Chemotherapy-IO across all relevant PD-L1 subgroups, integrating TMB and genomic analyses for the first time.

Our study highlights the critical nature of ongoing clinical trials prospectively evaluating the comparative effectiveness of anti-PD-(L)1 with and without chemotherapy (NCT03793179, NCT04547504). Our population included patients often not well-represented in randomized-controlled trials. These trials exclude patients with ECOG performance status of ≥2 and patients with active, untreated, or symptomatic brain metastases. Our study population comprised 15% with ECOG performance status ≥2 and 30% with baseline brain metastases.

In conclusion, we observed improvements in ORR and PFS associated with Chemotherapy-IO compared to IO, particularly in young, never-smokers. Never smoking status in the PD-L1 ≥50% subgroup was the only characteristic in which ORR and PFS improvement translated into a survival benefit. No genomic alterations favored overall survival with Chemotherapy-IO compared to IO. Our findings demonstrate that the addition of chemotherapy to PD-(L)1 blockade generally increases the probability of initial response, but leads to improved survival only among never-smokers in the PD-L1 ≥50% subgroup.

## Data Availability

Please see details in the manuscript.

## Acknowledgements

The authors wish to acknowledge Clare Wilhelm for editorial support. The authors wish to acknowledge the support of the Center Core grant (P30 CA008748), the Molecular Diagnostics Service in the Department of Pathology, and the Marie-Josee and Henry R. Kravis Center for Molecular Oncology.

## List of abbreviations

NSCLC: Non-small cell lung cancer
LUAD: Lung adenocarcinoma
IO: Anti-PD-(L)-1 monotherapy
Chemotherapy-IO: Combination anti-PD-(L)-1 with platinum-doublet chemotherapy
ORR: Overall response rate
PFS: Progression-free survival
OS: Overall survival
HR: Hazard ratio
95%CI: 95% confidence interval
NGS: Next-generation sequencing
PD-L1 TPS: PD-L1 tumor proportion score

## Supplemental Figure Legends

**Figure S1. Cohort assembly**. *A total of N=3777 patients treated with either Chemotherapy-IO or IO treated between 1/10/17 and 9/8/20*. Chemotherapy-IO; Combination PD(L)-1 blockade and platinum chemotherapy. IO; single agent PD(L)-1 blockade.

**Figure S2. Median PD-L1 and harmonized tumor mutational burden (TMB) between chemotherapy plus PD-(L)1 blockade (Chemotherapy-IO) vs single-agent PD-(L)1 blockade (IO) groups. A**. Box plots of tumor PD-L1 expression and **B**. Median TMB between Chemotherapy-IO and IO. ***p<0.001

**Figure S3. Outcomes to chemotherapy plus PD-(L)1 blockade (Chemotherapy-IO) vs single-agent PD-(L)1 blockade (IO) among patients with PD-L1 ≥90%. A**. Objective response rate, **B**. Progression-free survival and **C**. Overall survival. Chemotherapy-IO and IO. Chemotherapy-IO; Combination PD(L)-1 blockade and platinum chemotherapy. IO; single agent PD(L)-1 blockade.

**Figure S4. Comparative effectiveness of chemotherapy plus PD-(L)1 blockade (Chemotherapy-IO) vs single-agent PD-(L)1 blockade (IO) in patients with PD-L1 1-49%. A**. Objective response rate, **B**. Progression-free survival (PFS), and **C**. Overall survival (OS) among Chemotherapy-IO vs IO groups. **D**. PFS and **E**. OS analysis for Chemotherapy-IO vs IO adjusting for co-variates of interest among different subgroups. ECOG; Eastern Cooperative Oncology Group performance status. Median survival times presented with confidence intervals in brackets. HR; hazard ratio. 95%CI; 95% confidence intervals.

## Notes

**Competing interests:** JEC has consulted for: AstraZeneca, BMS, Genentech, Merck, Flame Biosciences, Regeneron-Sanofi, Guardant Health, Arcus Biosciences and also declared research funding to institution from: AstraZeneca, BMS, Genentech, Merck, Novartis. CMR has consulted with AbbVie, Amgen, Astra Zeneca, D2G, Daiichi Sankyo, Epizyme, Genentech/Roche, Ipsen, Jazz, Kowa, Lilly, Merck, and Syros. He serves on the scientific advisory boards of Bridge Medicines, Earli, and Harpoon Therapeutics. GJR reports institutional research support from Mirati, Lilly, Takeda, Merck, Roche, Pfizer, and Novartis. MGK reports honoraria from AstraZeneca, Pfizer, Merus, and BerGenBio. MDH reports grants from BMS; and personal fees from Achilles; Adagene; Adicet; Arcus; AstraZeneca; Blueprint; BMS; DaVolterra; Eli Lilly; Genentech/Roche; Genzyme/Sanofi; Janssen; Immunai; Instil Bio; Mana Therapeutics; Merck; Mirati; Natera; Pact Pharma; Shattuck Labs; and Regeneron; as well as equity options from Factorial, Immunai, Shattuck Labs, Arcus, and Avail Bio. A patent filed by Memorial Sloan Kettering related to the use of tumor mutational burden to predict response to immunotherapy (PCT/US2015/062208) is pending and licensed by PGDx. Subsequent to the completion of this work, MDH began as an employee (and equity holder) at AstraZeneca. MMA reports that he was a consultant to: Bristol-Myers Squibb, Merck, Genentech, AstraZeneca, Mirati, Novartis, Blueprint Medicine, Abbvie, Gritstone, NextCure, EMD Serono and received research funding (to institute) from: Bristol-Myers Squibb, Eli Lilly, Genentech, AstraZeneca, Amgen. AJS reports consulting/advising role to J&J, KSQ therapeutics, BMS, Merck, Enara Bio, Perceptive Advisors, Oppenheimer and Co, Umoja Biopharma, Legend Biotech, Iovance Biotherapeutics, Lyell Immunopharma and Heat Biologics, research funding from GSK (Inst), PACT pharma (Inst), Iovance Biotherapeutics (Inst), Achilles therapeutics (Inst), Merck (Inst), BMS (Inst), Harpoon Therapeutics (Inst) and Amgen (Inst). AJS reports consulting/advising role to J&J, KSQ therapeutics, BMS, Merck, Enara Bio, Perceptive Advisors, Oppenheimer and Co, Umoja Biopharma, Legend Biotech, Iovance Biotherapeutics, Lyell Immunopharma and Heat Biologics. Research funding: GSK (Inst), PACT pharma (Inst), Iovance Biotherapeutics (Inst), Achilles therapeutics (Inst), Merck (Inst), BMS (Inst), Harpoon Therapeutics (Inst) and Amgen (Inst).

### Competing Interest Statement

Please see manuscript for details.

### Author Declarations

This study was approved by Memorial Sloan Kettering Cancer Center and Dana-Farber Cancer Institute IRB.

### Summary of Updates

Manuscript, figures, and supplementary figures revised.

