## Supplemental Tables for "Efficacy of PD-(L)1 blockade monotherapy compared to PD-(L)1 blockade plus chemotherapy in first-line PD-L1-positive advanced lung adenocarcinomas: A cohort study"

Supplementary Material: Table of Contents

### **Supplemental Methods**

**Genomic sequencing – Memorial Sloan Kettering Cancer Center (MSK)**

Biopsies from patients treated at MSK underwent next-generation sequencing (NGS) using the MSK-IMPACT platform as previously described^34^. Briefly, DNA was extracted from tumors and patient-matched blood samples. Bar-coded libraries were generated and sequenced for targeted all exons and select introns of a custom gene panel of 341 (version1), 410 (version 2), or 468 (version 3) genes. Samples were run through a custom pipeline to identify somatic alterations, including mutations and copy number alterations. Tumor mutational burden was calculated as previously described^35^.

**Genomic sequencing – Dana-Farber Cancer Institute (DFCI)**

Targeted exome NGS (Profile) was carried out using the validated OncoPanel assay in the Center for Cancer Genome Discovery at the DFCI for 277 (POPv1), 302 (POPv2), or 447 (POPv3) cancer-associated genes. Variants were filtered to remove potential germline variants as previously published and annotated using Oncotractor as previously described. To remove additional germline noise, variants that were annotated as benign/likely benign in ClinVar or were present at a population maximum allele frequency of < 0.1% were excluded. Variants were retained in either case if they were annotated as confirmed somatic in at least two samples in COSMIC as previously described^36^.

**Harmonization of** **Tumor Mutation Burden**

Tumor mutational burden was calculated at MSK and at DFCI as previously described. To address differences in sequencing methodologies, we performed harmonization of TMB as previously done^15^. TMB distributions were harmonized by applying a normal transformation followed by standardization to z-scores, which enables integration of datasets derived from different sequencing panels^15^.

### **Table S1: PD-L1≥1%: Propensity score analysis for Chemo-IO vs. IO determining likelihood of receiving Chemo-IO vs. IO**

| **PD-L1≥1%: Propensity score analysis for Chemo/IO vs. IO determining**  **likelihood of receiving Chemo/IO vs. IO** | | | |
| --- | --- | --- | --- |
| **Characteristic** | **OR***^1^* | **95% CI***^1^* | **p-value** |
| >=65 vs. <65 | 0.77 | 0.54, 1.09 | 0.14 |
| ECOG 2-3 vs. 0-1 | 0.85 | 0.53, 1.36 | 0.5 |
| Current/Former Smoker vs. Never Smoker | 0.48 | 0.28, 0.79 | **0.004** |
| PD-L1 >=50 vs. 1-49 | 0.07 | 0.05, 0.10 | **<0.001** |
| Baseline liver metastases | 0.72 | 0.43, 1.18 | 0.2 |
| Baseline brain metastases | 0.85 | 0.57, 1.24 | 0.4 |
| *^1^* OR = Odds Ratio, CI = Confidence Interval | | | |

### **Table S2: PD-L1≥1%: Propensity-adjusted Cox model for PFS – Main effects model**

| **PD-L1 ≥1%: Propensity-adjusted Cox model for PFS - Main effects model** | | | |
| --- | --- | --- | --- |
| **Characteristic** | **HR***^1^* | **95% CI***^1^* | **p-value** |
| Chemo + IO vs. IO | 0.75 | 0.61, 0.92 | **0.005** |
| >=65 vs. <65 | 1.29 | 1.06, 1.58 | **0.013** |
| ECOG (2-3 vs. 0-1) | 1.44 | 1.09, 1.90 | **0.009** |
| Current/Former Smoker vs. Never Smoker | 0.88 | 0.64, 1.21 | 0.4 |
| PD-L1 >=50 vs. 1-49 | 0.74 | 0.61, 0.90 | **0.003** |
| Baseline liver metastases | 1.61 | 1.19, 2.17 | **0.002** |
| Baseline brain metastases | 1.05 | 0.83, 1.31 | 0.7 |
| *^1^* HR = Hazard Ratio, CI = Confidence Interval | | | |

### **Table S3: PD-L1 ≥1%: Propensity-adjusted Cox model for OS - Main effects model**

| **PD-L1 ≥1%: Propensity-adjusted Cox model for OS - Main effects model** | | | |
| --- | --- | --- | --- |
| **Characteristic** | **HR***^1^* | **95% CI***^1^* | **p-value** |
| Chemo + IO vs. IO | 1.05 | 0.83, 1.33 | 0.7 |
| >=65 vs. <65 | 1.44 | 1.14, 1.83 | **0.002** |
| ECOG (2-3 vs. 0-1) | 1.99 | 1.45, 2.74 | **<0.001** |
| Current/Former Smoker vs. Never Smoker | 1.29 | 0.93, 1.79 | 0.13 |
| PD-L1 >=50 vs. 1-49 | 0.79 | 0.62, 0.99 | **0.044** |
| Baseline liver metastases | 1.48 | 1.05, 2.09 | **0.025** |
| Baseline brain metastases | 0.97 | 0.74, 1.27 | 0.8 |
| *^1^* HR = Hazard Ratio, CI = Confidence Interval | | | |

### **Table S4: PD-L1 ≥1%: Propensity-adjusted Cox model for PFS – Treatment interaction model**

| Propensity-adjusted Cox model for PFS (All patients) | | | |
| --- | --- | --- | --- |
| **Characteristic** | **HR***^1^* | **95% CI***^1^* | **p-value** |
| Chemo + IO vs. IO | 0.38 | 0.17, 0.86 | **0.020** |
| >=65 vs. <65 | 1.06 | 0.81, 1.39 | 0.7 |
| ECOG (2-3 vs. 0-1) | 1.46 | 1.05, 2.02 | **0.025** |
| Current/Former Smoker vs. Never Smoker | 0.66 | 0.37, 1.17 | 0.2 |
| PD-L1 >=50 vs. 1-49 | 0.79 | 0.59, 1.06 | 0.12 |
| Baseline liver metastases | 1.61 | 1.13, 2.30 | **0.009** |
| Baseline brain metastases | 1.02 | 0.76, 1.36 | 0.9 |
| Chemo + IO vs. IO * >=65 vs. <65 | 1.49 | 0.99, 2.25 | 0.055 |
| Chemo + IO vs. IO * ECOG (2-3 vs. 0-1) | 0.91 | 0.51, 1.63 | 0.8 |
| Chemo + IO vs. IO * Current/Former Smoker vs. Never Smoker | 1.73 | 0.89, 3.35 | 0.11 |
| Chemo + IO vs. IO * PD-L1 >=50 vs. 1-49 | 0.90 | 0.60, 1.34 | 0.6 |
| Chemo + IO vs. IO * Baseline liver metastases | 1.05 | 0.57, 1.92 | 0.9 |
| Chemo + IO vs. IO * Baseline brain metastases | 1.03 | 0.66, 1.63 | 0.9 |
| *^1^* HR = Hazard Ratio, CI = Confidence Interval | | | |

### **Table S5: PD-L1 ≥1%: Propensity-adjusted Cox model for OS – Treatment interaction model**

| Propensity-adjusted Cox model for OS (All patients) | | | |
| --- | --- | --- | --- |
| **Characteristic** | **HR***^1^* | **95% CI***^1^* | **p-value** |
| Chemo + IO vs. IO | 0.92 | 0.42, 2.00 | 0.8 |
| >=65 vs. <65 | 1.40 | 1.04, 1.89 | **0.029** |
| ECOG (2-3 vs. 0-1) | 2.27 | 1.58, 3.26 | **<0.001** |
| Current/Former Smoker vs. Never Smoker | 1.20 | 0.72, 2.00 | 0.5 |
| PD-L1 >=50 vs. 1-49 | 0.82 | 0.60, 1.13 | 0.2 |
| Baseline liver metastases | 1.38 | 0.93, 2.05 | 0.11 |
| Baseline brain metastases | 0.91 | 0.66, 1.27 | 0.6 |
| Chemo + IO vs. IO * >=65 vs. <65 | 1.07 | 0.66, 1.73 | 0.8 |
| Chemo + IO vs. IO * ECOG (2-3 vs. 0-1) | 0.75 | 0.39, 1.45 | 0.4 |
| Chemo + IO vs. IO * Current/Former Smoker vs. Never Smoker | 1.17 | 0.61, 2.25 | 0.6 |
| Chemo + IO vs. IO * PD-L1 >=50 vs. 1-49 | 0.92 | 0.58, 1.48 | 0.7 |
| Chemo + IO vs. IO * Baseline liver metastases | 1.16 | 0.57, 2.36 | 0.7 |
| Chemo + IO vs. IO * Baseline brain metastases | 1.14 | 0.66, 1.96 | 0.6 |
| *^1^* HR = Hazard Ratio, CI = Confidence Interval | | | |

### **Table S6: PD-L1≥50%: Propensity score analysis for Chemo-IO vs. IO determining likelihood of receiving Chemo-IO vs. IO**

| **PD-L1≥50%: Propensity score analysis for Chemo/IO vs. IO determining**  **likelihood of receiving Chemo/IO vs. IO** | | | |
| --- | --- | --- | --- |
| **Characteristic** | **OR***^1^* | **95% CI***^1^* | **p-value** |
| >=65 vs. <65 | 0.58 | 0.38, 0.90 | **0.015** |
| ECOG 2-3 vs. 0-1 | 0.94 | 0.50, 1.68 | 0.8 |
| Current/Former Smoker vs. Never Smoker | 0.52 | 0.28, 1.01 | **0.045** |
| Baseline liver metastases | 0.72 | 0.36, 1.34 | 0.3 |
| Baseline brain metastases | 0.84 | 0.51, 1.35 | 0.5 |
| *^1^* OR = Odds Ratio, CI = Confidence Interval | | | |

### **Table S7: PD-L1 ≥50%: Propensity-adjusted Cox model for PFS - Main effects model**

| **PD-L1 ≥50%: Propensity-adjusted Cox model for PFS - Main effects model** | | | |
| --- | --- | --- | --- |
| **Characteristic** | **HR***^1^* | **95% CI***^1^* | **p-value** |
| Chemo + IO vs. IO | 0.74 | 0.56, 0.97 | **0.032** |
| >=65 vs. <65 | 1.34 | 1.03, 1.75 | **0.027** |
| ECOG (2-3 vs. 0-1) | 1.38 | 0.93, 2.04 | 0.11 |
| Current/Former Smoker vs. Never Smoker | 0.91 | 0.63, 1.32 | 0.6 |
| Baseline liver metastases | 1.70 | 1.17, 2.48 | **0.005** |
| Baseline brain metastases | 0.95 | 0.72, 1.27 | 0.7 |
| *^1^* HR = Hazard Ratio, CI = Confidence Interval | | | |

### **Table S8: PD-L1 ≥50%: Propensity-adjusted Cox model for OS - Main effects model**

| **PD-L1 ≥50%: Propensity-adjusted Cox model for OS - Main effects model** | | | |
| --- | --- | --- | --- |
| **Characteristic** | **HR***^1^* | **95% CI***^1^* | **p-value** |
| Chemo + IO vs. IO | 1.02 | 0.73, 1.43 | >0.9 |
| >=65 vs. <65 | 1.51 | 1.10, 2.07 | **0.011** |
| ECOG (2-3 vs. 0-1) | 2.00 | 1.27, 3.14 | **0.003** |
| Current/Former Smoker vs. Never Smoker | 1.24 | 0.78, 1.98 | 0.4 |
| Baseline liver metastases | 1.66 | 1.08, 2.56 | **0.020** |
| Baseline brain metastases | 0.88 | 0.61, 1.25 | 0.5 |
| *^1^* HR = Hazard Ratio, CI = Confidence Interval | | | |

| Propensity-adjusted Cox model for PFS | | | |
| --- | --- | --- | --- |
| **Characteristic** | **HR***^1^* | **95% CI***^1^* | **p-value** |
| Chemo + IO vs. IO | 0.21 | 0.09, 0.49 | **<0.001** |
| >=65 vs. <65 | 1.03 | 0.80, 1.32 | 0.8 |
| ECOG (2-3 vs. 0-1) | 1.35 | 0.97, 1.89 | 0.073 |
| Current/Former Smoker vs. Never Smoker | 0.57 | 0.37, 0.87 | **0.009** |
| Baseline liver metastases | 1.58 | 1.14, 2.20 | **0.007** |
| Baseline brain metastases | 0.99 | 0.76, 1.27 | >0.9 |
| Chemo + IO vs. IO * >=65 vs. <65 | 1.80 | 1.05, 3.09 | **0.032** |
| Chemo + IO vs. IO * ECOG (2-3 vs. 0-1) | 0.93 | 0.39, 2.24 | 0.9 |
| Chemo + IO vs. IO * Current/Former Smoker vs. Never Smoker | 2.61 | 1.27, 5.35 | **0.009** |
| Chemo + IO vs. IO * Baseline liver metastases | 1.23 | 0.56, 2.71 | 0.6 |
| Chemo + IO vs. IO * Baseline brain metastases | 0.88 | 0.48, 1.62 | 0.7 |
| *^1^* HR = Hazard Ratio, CI = Confidence Interval | | | |

### **Table S9: PD-L1 ≥50%: Propensity-adjusted Cox model for PFS – Treatment interaction model**

### **Table S10: PD-L1 ≥50%: Propensity-adjusted Cox model for OS – Treatment interaction model**

| Propensity-adjusted Cox model for OS | | | |
| --- | --- | --- | --- |
| **Characteristic** | **HR***^1^* | **95% CI***^1^* | **p-value** |
| Chemo + IO vs. IO | 0.36 | 0.12, 1.10 | 0.072 |
| >=65 vs. <65 | 1.37 | 1.03, 1.82 | **0.032** |
| ECOG (2-3 vs. 0-1) | 2.06 | 1.42, 2.98 | **<0.001** |
| Current/Former Smoker vs. Never Smoker | 0.78 | 0.50, 1.21 | 0.3 |
| Baseline liver metastases | 1.66 | 1.22, 2.27 | **0.001** |
| Baseline brain metastases | 0.96 | 0.72, 1.29 | 0.8 |
| Chemo + IO vs. IO * >=65 vs. <65 | 1.28 | 0.66, 2.48 | 0.5 |
| Chemo + IO vs. IO * ECOG (2-3 vs. 0-1) | 0.89 | 0.33, 2.39 | 0.8 |
| Chemo + IO vs. IO * Current/Former Smoker vs. Never Smoker | 2.93 | 1.08, 7.92 | **0.034** |
| Chemo + IO vs. IO * Baseline liver metastases | 0.99 | 0.37, 2.67 | >0.9 |
| Chemo + IO vs. IO * Baseline brain metastases | 0.78 | 0.36, 1.72 | 0.5 |
| *^1^* HR = Hazard Ratio, CI = Confidence Interval | | | |

### **Table S11: PD-L1 1-49%: Propensity score analysis for Chemo-IO vs. IO determining likelihood of receiving Chemo-IO vs. IO**

| **PD-L1 1-49%: Propensity score analysis for Chemo/IO vs. IO determining**  **likelihood of receiving Chemo/IO vs. IO** | | | |
| --- | --- | --- | --- |
| **Characteristic** | **OR***^1^* | **95% CI***^1^* | **p-value** |
| >=65 vs. <65 | 1.23 | 0.70, 2.14 | 0.5 |
| ECOG 2-3 vs. 0-1 | 0.73 | 0.36, 1.57 | 0.4 |
| Current/Former Smoker vs. Never Smoker | 0.37 | 0.12, 0.90 | **0.044** |
| Baseline liver metastases | 0.76 | 0.35, 1.74 | 0.5 |
| Baseline brain metastases | 0.94 | 0.50, 1.80 | 0.8 |
| *^1^* OR = Odds Ratio, CI = Confidence Interval | | | |

### **Table S12: PD-L1 1-49%: Propensity-adjusted Cox model for PFS - Main effects model**

| **PD-L1 1-49%: Propensity-adjusted Cox model for PFS - Main effects model** | | | |
| --- | --- | --- | --- |
| **Characteristic** | **HR***^1^* | **95% CI***^1^* | **p-value** |
| Chemo + IO vs. IO | 0.77 | 0.56, 1.07 | 0.12 |
| >=65 vs. <65 | 1.26 | 0.92, 1.73 | 0.14 |
| ECOG (2-3 vs. 0-1) | 1.53 | 1.05, 2.24 | **0.027** |
| Current/Former Smoker vs. Never Smoker | 0.84 | 0.48, 1.46 | 0.5 |
| Baseline liver metastases | 1.38 | 0.83, 2.28 | 0.2 |
| Baseline brain metastases | 1.30 | 0.90, 1.87 | 0.2 |
| *^1^* HR = Hazard Ratio, CI = Confidence Interval | | | |

### **Table S13: PD-L1 1-49%: Propensity-adjusted Cox model for OS - Main effects model**

| **PD-L1 1-49%: Propensity-adjusted Cox model for OS - Main effects model** | | | |
| --- | --- | --- | --- |
| **Characteristic** | **HR***^1^* | **95% CI***^1^* | **p-value** |
| Chemo + IO vs. IO | 1.10 | 0.78, 1.54 | 0.6 |
| >=65 vs. <65 | 1.41 | 0.99, 2.00 | 0.055 |
| ECOG (2-3 vs. 0-1) | 2.14 | 1.45, 3.18 | **<0.001** |
| Current/Former Smoker vs. Never Smoker | 1.49 | 0.90, 2.46 | 0.12 |
| Baseline liver metastases | 1.16 | 0.66, 2.02 | 0.6 |
| Baseline brain metastases | 1.22 | 0.81, 1.83 | 0.3 |
| *^1^* HR = Hazard Ratio, CI = Confidence Interval | | | |

| Propensity-adjusted Cox model for PFS | | | |
| --- | --- | --- | --- |
| **Characteristic** | **HR***^1^* | **95% CI***^1^* | **p-value** |
| Chemo + IO vs. IO | 0.60 | 0.15, 2.36 | 0.5 |
| >=65 vs. <65 | 1.22 | 0.69, 2.18 | 0.5 |
| ECOG (2-3 vs. 0-1) | 1.51 | 0.67, 3.40 | 0.3 |
| Current/Former Smoker vs. Never Smoker | 0.72 | 0.20, 2.54 | 0.6 |
| Baseline liver metastases | 1.49 | 0.58, 3.78 | 0.4 |
| Baseline brain metastases | 1.28 | 0.63, 2.58 | 0.5 |
| Chemo + IO vs. IO * >=65 vs. <65 | 1.04 | 0.55, 1.98 | 0.9 |
| Chemo + IO vs. IO * ECOG (2-3 vs. 0-1) | 0.98 | 0.41, 2.36 | >0.9 |
| Chemo + IO vs. IO * Current/Former Smoker vs. Never Smoker | 1.32 | 0.36, 4.83 | 0.7 |
| Chemo + IO vs. IO * Baseline liver metastases | 0.89 | 0.32, 2.44 | 0.8 |
| Chemo + IO vs. IO * Baseline brain metastases | 1.04 | 0.48, 2.25 | >0.9 |
| *^1^* HR = Hazard Ratio, CI = Confidence Interval | | | |

### **Table S14: PD-L1 1-49%: Propensity-adjusted Cox model for PFS – Treatment interaction model**

| Propensity-adjusted Cox model for OS | | | |
| --- | --- | --- | --- |
| **Characteristic** | **HR***^1^* | **95% CI***^1^* | **p-value** |
| Chemo + IO vs. IO | 2.74 | 0.87, 8.64 | 0.084 |
| >=65 vs. <65 | 1.65 | 0.85, 3.23 | 0.14 |
| ECOG (2-3 vs. 0-1) | 3.11 | 1.43, 6.77 | **0.004** |
| Current/Former Smoker vs. Never Smoker | 2.55 | 1.01, 6.43 | **0.047** |
| Baseline liver metastases | 0.92 | 0.33, 2.53 | 0.9 |
| Baseline brain metastases | 1.09 | 0.49, 2.41 | 0.8 |
| Chemo + IO vs. IO * >=65 vs. <65 | 0.76 | 0.36, 1.60 | 0.5 |
| Chemo + IO vs. IO * ECOG (2-3 vs. 0-1) | 0.48 | 0.19, 1.17 | 0.11 |
| Chemo + IO vs. IO * Current/Former Smoker vs. Never Smoker | 0.43 | 0.16, 1.18 | 0.10 |
| Chemo + IO vs. IO * Baseline liver metastases | 1.62 | 0.53, 4.96 | 0.4 |
| Chemo + IO vs. IO * Baseline brain metastases | 1.41 | 0.59, 3.37 | 0.4 |
| *^1^* HR = Hazard Ratio, CI = Confidence Interval | | | |

### **Table S15: PD-L1 1-49%: Propensity-adjusted Cox model for OS – Treatment interaction model**

### **Table S16: Baseline characteristics for genomic analysis cohort**

| **Characteristic** | **Overall, N = 579*^1^*** | **Chemo/IO, N = 263*^1^*** | **IO, N = 316*^1^*** | **p-value*^2^*** |
| --- | --- | --- | --- | --- |
| **Site** |  |  |  | 0.2 |
| DFCI | 319 (55%) | 138 (52%) | 181 (57%) |  |
| MSK | 260 (45%) | 125 (48%) | 135 (43%) |  |
| **Age** | 67 (60, 75) | 66 (59, 73) | 68 (60, 76) | **0.008** |
| **Sex** |  |  |  | 0.7 |
| Female | 331 (57%) | 148 (56%) | 183 (58%) |  |
| Male | 248 (43%) | 115 (44%) | 133 (42%) |  |
| **ECOG** |  |  |  | 0.3 |
| < 2 | 528 (93%) | 231 (92%) | 297 (94%) |  |
| >= 2 | 38 (6.7%) | 20 (8.0%) | 18 (5.7%) |  |
| Unknown | 13 | 12 | 1 |  |
| **Pack yrs** | 28 (10, 44) | 25 (7, 40) | 30 (15, 45) | **0.032** |
| Unknown | 27 | 20 | 7 |  |
| **Smoking Status** |  |  |  | **0.002** |
| Current/Former | 508 (88%) | 218 (83%) | 290 (92%) |  |
| Never | 70 (12%) | 44 (17%) | 26 (8.2%) |  |
| Unknown | 1 | 1 | 0 |  |
| **Baseline liver metastases** | 81 (14%) | 34 (13%) | 47 (15%) | 0.5 |
| **Baseline brain metastases** | 166 (29%) | 68 (26%) | 98 (31%) | 0.2 |
| Unknown | 2 | 2 | 0 |  |
| **PD-L1 Status** |  |  |  | **<0.001** |
| PD-L1 1-49 | 243 (42%) | 195 (74%) | 48 (15%) |  |
| PD-L1 >=50 | 336 (58%) | 68 (26%) | 268 (85%) |  |
| **Harmonized TMB score** | 0.09 (-0.68, 0.65) | -0.05 (-0.73, 0.55) | 0.09 (-0.53, 0.66) | **0.023** |
| ***SMARCA4*** | 71 (12%) | 31 (12%) | 40 (13%) | 0.8 |
| ***STK11*** | 92 (16%) | 55 (21%) | 37 (12%) | **0.003** |
| ***KRAS*** | 301 (52%) | 136 (52%) | 165 (52%) | >0.9 |
| ***KEAP1*** | 116 (20%) | 57 (22%) | 59 (19%) | 0.4 |
| ***TP53*** | 351 (61%) | 152 (58%) | 199 (63%) | 0.2 |
| *^1^* n (%); Median (IQR) | | | | |
| *^2^* Pearson's Chi-squared test; Wilcoxon rank sum test | | | | |

| Propensity-adjusted Cox model for PFS | | | |
| --- | --- | --- | --- |
| **Characteristic** | **HR***^1^* | **95% CI***^1^* | **p-value** |
| Chemo + IO vs. IO | 0.44 | 0.17, 1.15 | 0.093 |
| >=65 vs. <65 | 1.14 | 0.83, 1.57 | 0.4 |
| ECOG (2-3 vs. 0-1) | 1.61 | 0.87, 2.99 | 0.13 |
| Current/Former Smoker vs. Never Smoker | 0.74 | 0.36, 1.51 | 0.4 |
| PD-L1 >=50 vs. 1-49 | 0.67 | 0.48, 0.92 | **0.013** |
| Baseline liver metastases | 1.17 | 0.75, 1.82 | 0.5 |
| Baseline brain metastases | 1.05 | 0.74, 1.49 | 0.8 |
| Chemo + IO vs. IO * >=65 vs. <65 | 1.26 | 0.76, 2.08 | 0.4 |
| Chemo + IO vs. IO * ECOG (2-3 vs. 0-1) | 0.81 | 0.31, 2.15 | 0.7 |
| Chemo + IO vs. IO * Current/Former Smoker vs. Never Smoker | 1.49 | 0.64, 3.44 | 0.4 |
| Chemo + IO vs. IO * PD-L1 >=50 vs. 1-49 | 1.05 | 0.65, 1.70 | 0.8 |
| Chemo + IO vs. IO * Baseline liver metastases | 1.46 | 0.71, 3.00 | 0.3 |
| Chemo + IO vs. IO * Baseline brain metastases | 0.75 | 0.42, 1.34 | 0.3 |
| *^1^* HR = Hazard Ratio, CI = Confidence Interval | | | |

### **Table S17: PD-L1 ≥1% Propensity-adjusted Cox model for PFS – Treatment interaction model**

| Propensity-adjusted Cox model for OS | | | |
| --- | --- | --- | --- |
| **Characteristic** | **HR***^1^* | **95% CI***^1^* | **p-value** |
| Chemo + IO vs. IO | 0.74 | 0.28, 1.95 | 0.5 |
| >=65 vs. <65 | 1.56 | 1.07, 2.28 | **0.021** |
| ECOG (2-3 vs. 0-1) | 2.07 | 1.16, 3.69 | **0.013** |
| Current/Former Smoker vs. Never Smoker | 1.16 | 0.57, 2.35 | 0.7 |
| PD-L1 >=50 vs. 1-49 | 0.59 | 0.41, 0.85 | **0.005** |
| Baseline liver metastases | 1.07 | 0.66, 1.73 | 0.8 |
| Baseline brain metastases | 0.95 | 0.63, 1.44 | 0.8 |
| Chemo + IO vs. IO * >=65 vs. <65 | 0.91 | 0.51, 1.63 | 0.8 |
| Chemo + IO vs. IO * ECOG (2-3 vs. 0-1) | 0.69 | 0.26, 1.85 | 0.5 |
| Chemo + IO vs. IO * Current/Former Smoker vs. Never Smoker | 1.25 | 0.52, 3.01 | 0.6 |
| Chemo + IO vs. IO * PD-L1 >=50 vs. 1-49 | 1.52 | 0.87, 2.65 | 0.14 |
| Chemo + IO vs. IO * Baseline liver metastases | 1.49 | 0.69, 3.24 | 0.3 |
| Chemo + IO vs. IO * Baseline brain metastases | 1.02 | 0.53, 1.98 | >0.9 |
| *^1^* HR = Hazard Ratio, CI = Confidence Interval | | | |

### **Table S18: PD-L1 ≥1% Propensity-adjusted Cox model for OS – Treatment interaction model**
