## Supplementary figures and images for "Efficacy of PD-(L)1 blockade monotherapy compared to PD-(L)1 blockade plus chemotherapy in first-line PD-L1-positive advanced lung adenocarcinomas: A cohort study"

### FigureS1

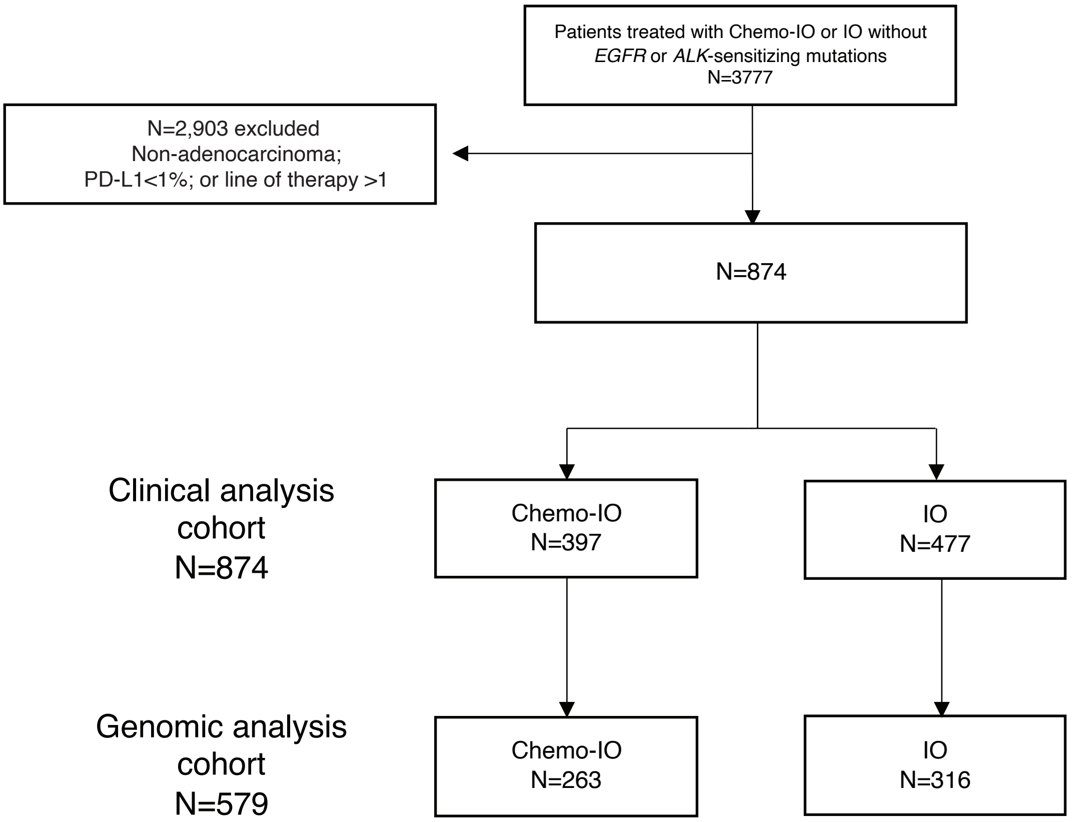

### FigureS2

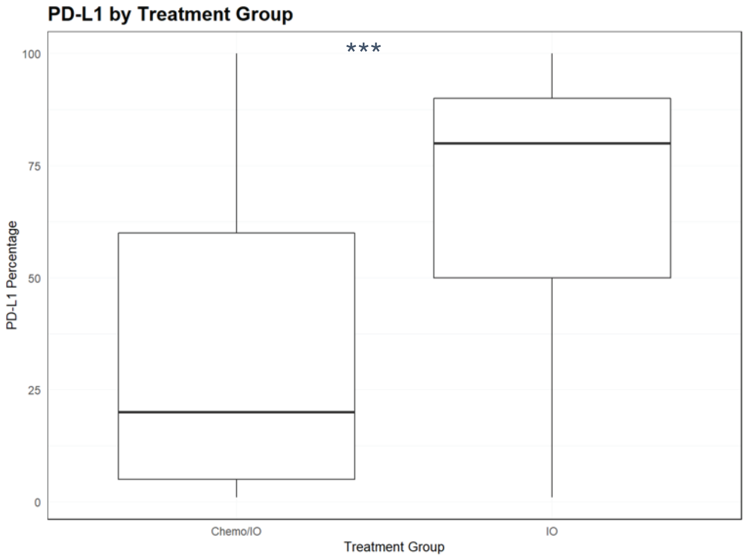

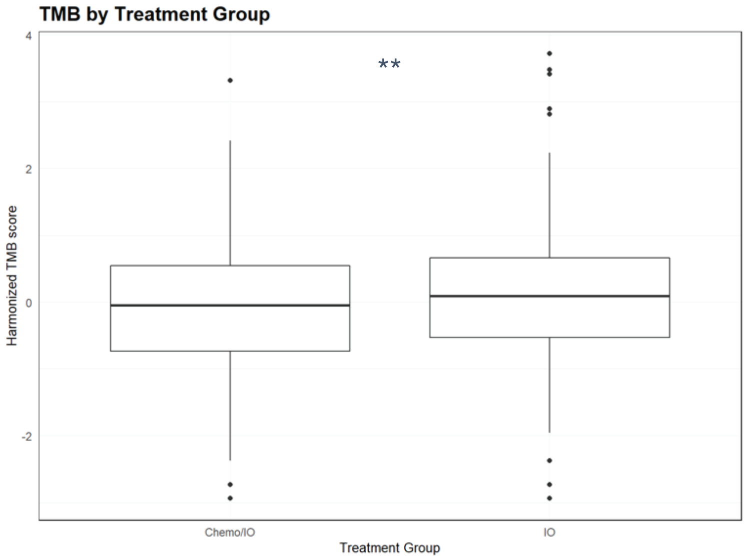

### FigureS3

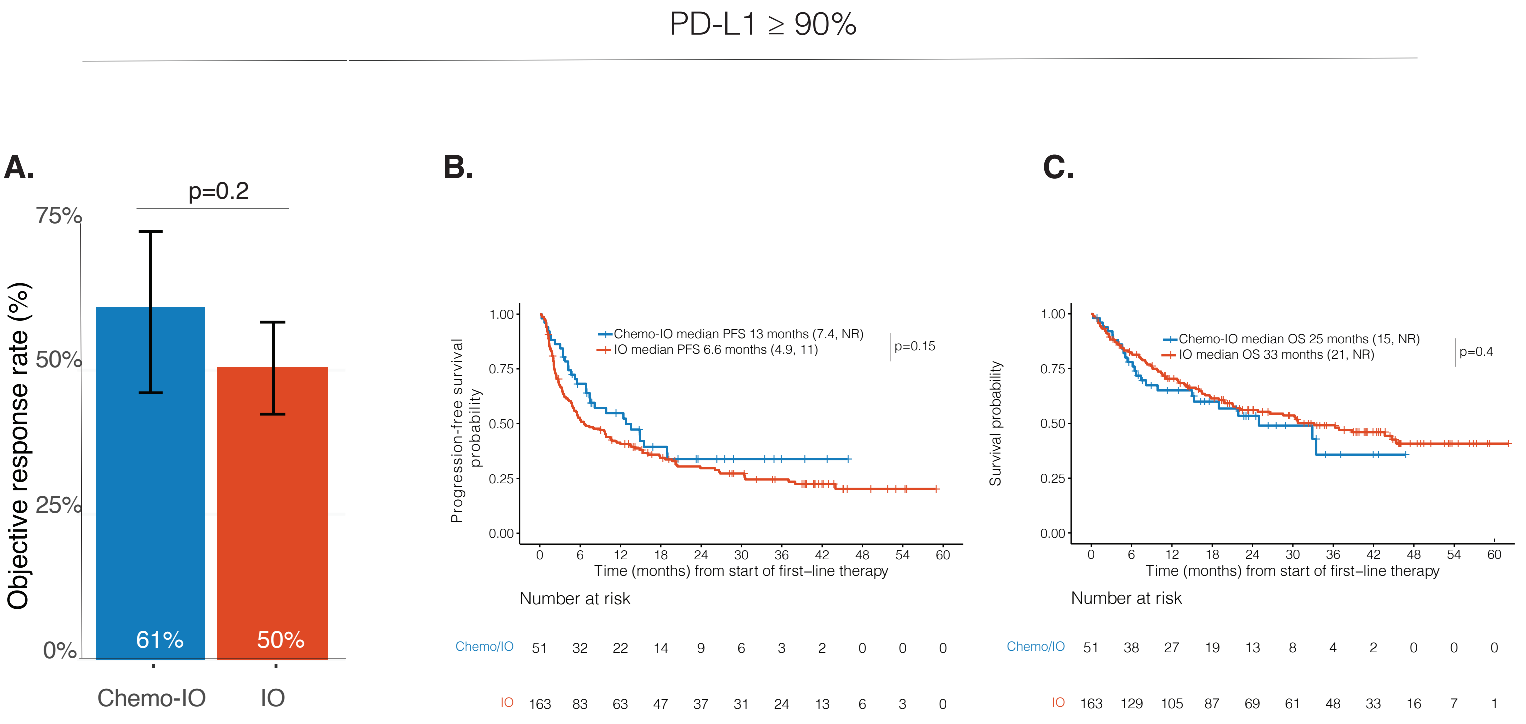

### FigureS4

**Figure S4**

PD-L1 1-49%

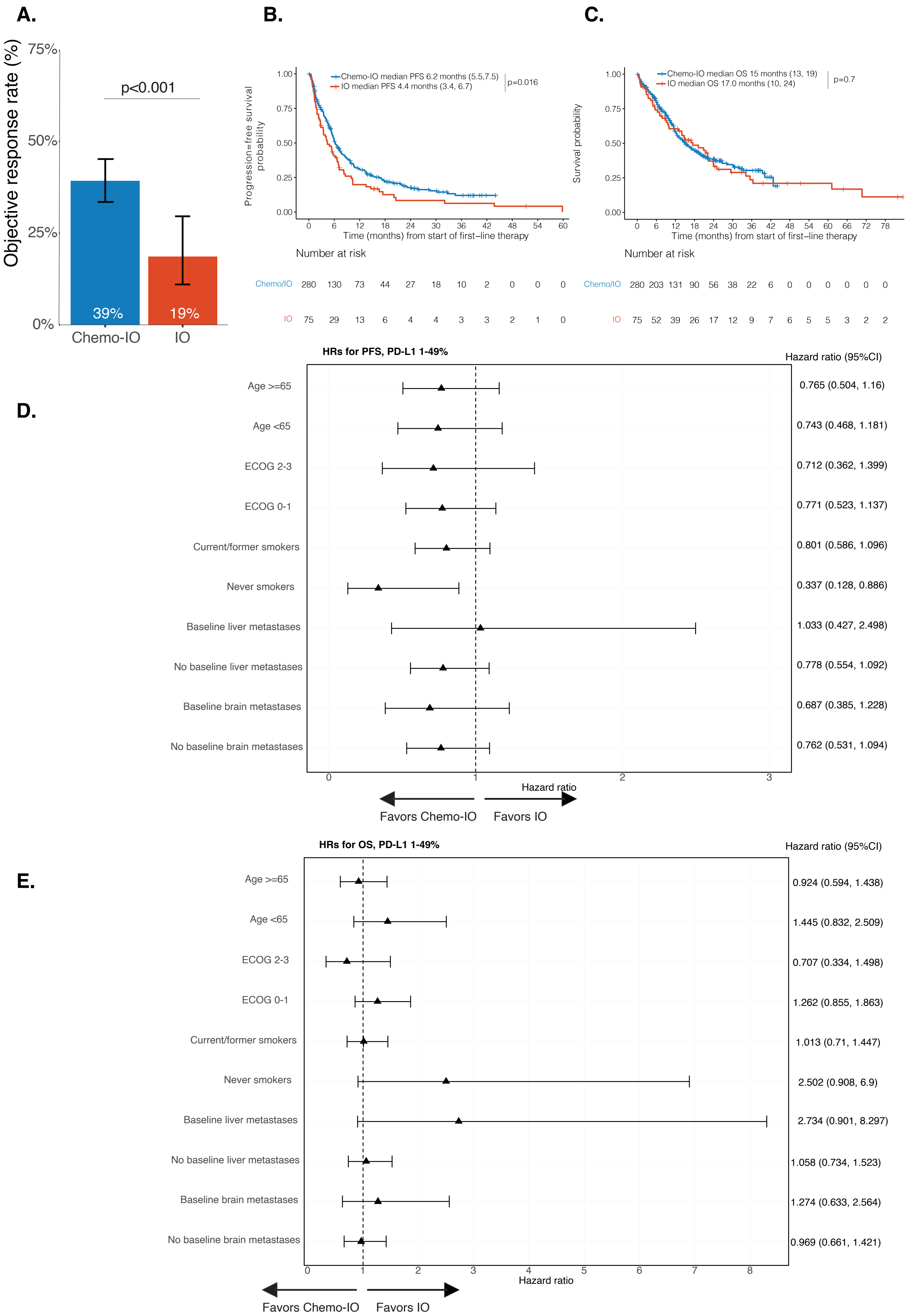
